# Automated characterisation of cerebral microbleeds using their size and spatial distribution on brain MR images

**DOI:** 10.1101/2025.01.10.25320354

**Authors:** Vaanathi Sundaresan, Giovanna Zamboni, Robert A Dineen, Dorothee P Auer, Stamatios N Sotiropoulos, Nikola Sprigg, Mark Jenkinson, Ludovica Griffanti

## Abstract

Cerebral microbleeds (CMBs) are small, hypointense hemosiderin deposits in the brain measuring around 2-10 mm in diameter. As one of the important biomarkers of small vessel disease, they have been associated with various neurodegenerative and cerebrovascular diseases. Hence, automated detection, and subsequent extraction of clinically useful metrics (e.g., size and spatial distribution) from CMBs are essential for investigating their clinical impact, especially in large-scale studies. While some work has been done for CMB segmentation, extraction of clinically relevant information is not yet explored. Herein, we propose the first automated method to characterise CMBs using their size and spatial distribution i.e., CMB count in three regions (and their sub-structures) used in microbleed anatomical rating scale (MARS): infratentorial, deep and lobar. Our method uses structural atlases of the brain for determining individual regions. On an intracerebral haemorrhage study dataset, we achieve a mean absolute error of 2mm for size estimation and an overall accuracy *>*90% for automated rating. The code and the atlas of MARS regions in MNI space are publicly available.

**Key points:**

- We present a method to automatically characterise cerebral microbleeds (size and location).
- The method achieved a mean absolute error of 2 mm for size estimation.
- Automated rating for infratentorial, deep and lobar regions achieved overall accuracy >90%
- We made the code and atlas of MARS regions publicly available.

## 1 Background

Cerebral microbleeds (CMBs) are hemosiderin deposits in the brain, mainly associated with cerebrovascular disease, Alzheimer’s disease and cerebral amyloid angiopathy (CAA) [1]. They appear as small, circular well-defined hypointense lesions of diameter 2-10mm on susceptibility weighted images (SWI). Automated CMB detection algorithms typically generate binary lesion maps of CMBs that are useful in determining the performance of the methods. However, for these methods to be translated into clinically deployable tools, their characterisation in terms of clinically useful metrics (e.g., size, count and spatial distribution) is a crucial step [2, 3]. This would provide the results to clinicians in easily interpretable terms in a time-efficient manner. In clinical settings, CMBs are often reported using rating scales, based on count and distribution of CMBs. Two manual rating scales have been proposed: microbleed anatomical rating scale (MARS) [3] and the brain observer microbleed scale (BOMBS) [4]. Among the two, while BOMBS provides the CMB count in all cerebral lobes combined, MARS provides CMB counts in individual lobes [3]. Obtaining such information also helps in better understanding of the clinical impact of CMBs. For instance, subcortical (deep) CMB are associated with cerebrovascular diseases, while lobar CMBs are associated with CAA [5]. In this work, we propose the first automated method to characterise CMBs from their binary maps by determining their sizes and their spatial distribution. We also show that obtaining the spatial distribution of CMBs could also be used to analyse the CMB detection performance better, by providing the detection statistics at region-level, in addition to providing clinically useful metrics for further population-level analysis.

## 2 Methods

### 2.1 Datasets and preprocessing

We used a subset of MRI data used as part of Tranexamic acid for IntraCerebral Haemorrhage2 (TICH2) trial [6, 7], consisting of 50 SWI (for demographic details and MRI acquisition parameters, refer to [7]). Out of 50 subjects, 25 subjects have CMBs. Manual segmentations and MARS ratings for CMBs are available for all 25 subjects with CMBs. Total number of CMBs: 185, mean: 7.5 ± 12.6 CMBs/subject, median: 5 CMBs/subject. For evaluation purposes, we included in our experiments all CMBs that were labelled as either ‘definite’ or ‘possible’. We reoriented the images to match the orientation of standard MNI space and skull stripped them using FSL BET [8], followed by bias field correction using FSL FAST [9]. For getting binary maps for CMB detection, we detected initial candidates using a shallow 3-layer U-Net [10], and then discriminated between true CMBs and mimics using the student model used in [11].

### 2.2 Automated characterisation of CMBs

The aim is to obtain the size of CMBs from the binary maps and use structural brain atlases to define the regions for obtaining region-wise counts of CMBs (shown in Fig. 1)

**Fig. 1.**
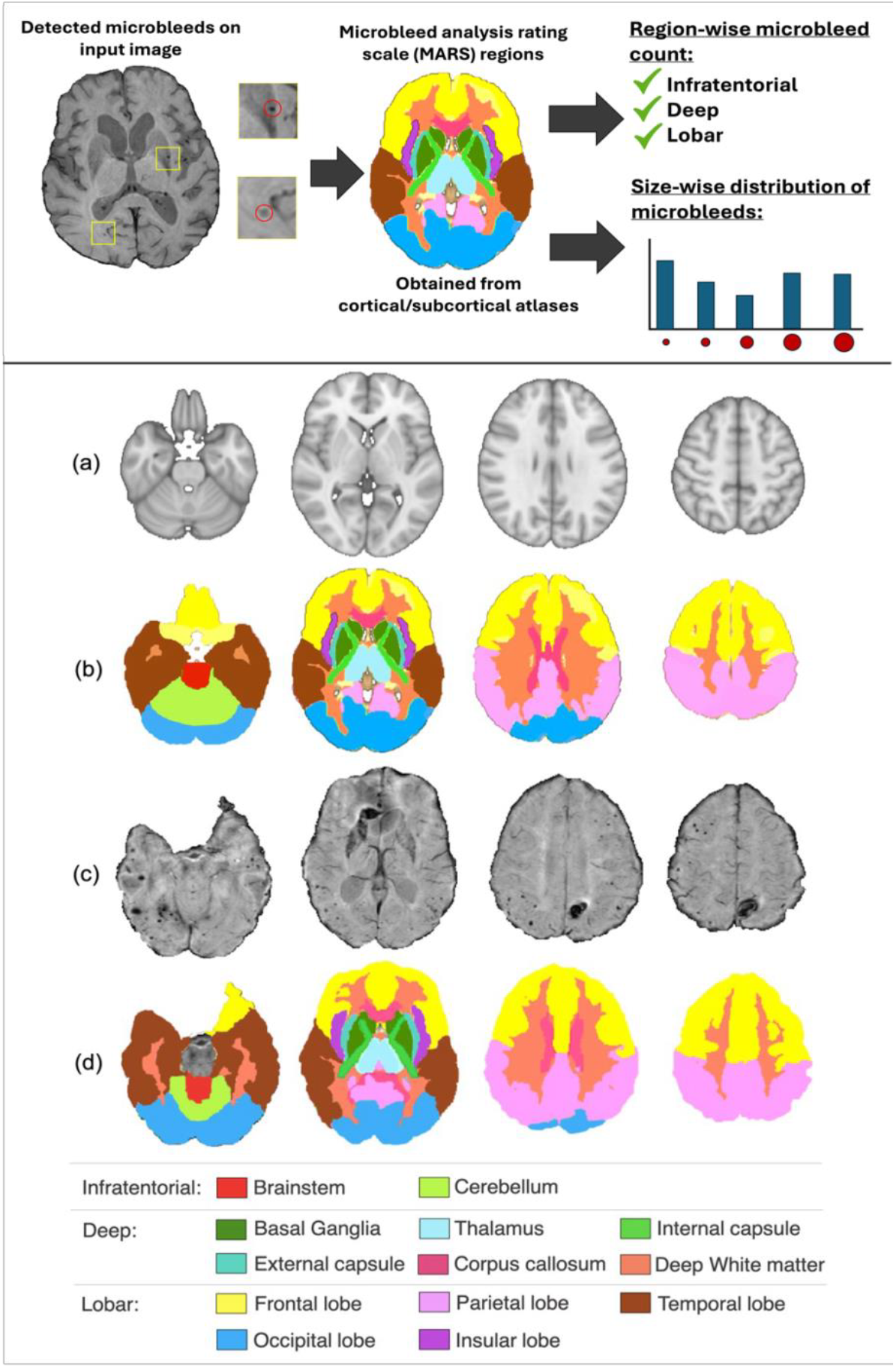
Proposed pipeline for region-wise characterisation of CMBs and MARS rating regions from the structural atlases. Top panel: Determination of CMB sizes and their spatial distribution from binary maps; bottom panel: (a) MNI T1-weighted structural image, (b) regions for MARS rating formed by using atlases in the MNI space, (c) SWI in the subject space, (d) MARS rating regions formed by registering atlases to the subject space. Legend for various structures shown at the bottom panel.

#### Estimation of CMB size

Within the bounding box around each detected CMB, we determined the major-axis length (the longest diameter of an ellipsoid within the bounding box, in voxels) of the individual CMBs and later scaled them with voxel dimensions to obtain the final diameters in mm.

#### Determination of spatial distribution of CMBs

We performed two steps: (1) extracting the regions (e.g., infratentorial) and structures within regions (e.g., brainstem) used in the MARS scale from MRI atlases and (2) counting the number of CMBs within each structure/region. For the first step, Table 1 reports the structures considered in the MARS scale and the atlases in the MNI space used for obtaining individual structures. For non-binary, probabilistic atlases we thresholded (values in Table 1) to obtain the final binary masks for individual structures. We later registered the structures from the MNI space to the subject-space using linear registration with 12 degrees of freedom with FSL-FLIRT [12] and thresholded them at 0.5 to obtain binary masks in the single-subject space. We performed a morphological dilation operation (using a structuring element with the size of 3 voxels) on the cortical lobes mask. This increased the extent of cortex to include the gray-white matter junctions as ‘lobar’ region. Also, deep structures such as internal/external capsules and corpus callosum were excluded from the white matter mask since they were considered separately. The implementation in Python and MARS region atlas are available at https://github.com/v-sundaresan/microbleed-size-counting.

Fig. 1 shows sample regions from the structural atlases in the MNI space and in the subject space. We then obtained the count of CMB centroids to obtain the structure-wise count of CMBs and aggregated the number of CMBs to provide region-wise CMB count.

### 2.3 Experimental setup and performance metrics

For CMB size estimation, we determined absolute error (AE) between sizes of the detected CMBs and of the manually segmented CMBs. For evaluation of automated MARS ratings using the detected CMBs, we compared the region-wise CMB counts from the predicted true positive (TP) CMBs to the manually rated MARS sub-scores. For region-wise count of CMBs, we used region-wise sensitivity, region-wise precision and region-wise F1-measure with respect to the manual ratings. We studied two different aspects: (i) changes in centroid locations due to variations in CMB outlines due to under/over-segmentation, leading to counting errors (from confusion matrices in Fig. 2(b)), and (ii) detection errors in case of false negatives (FNs) (shown in Fig. 2(c)).

**Fig. 2.**
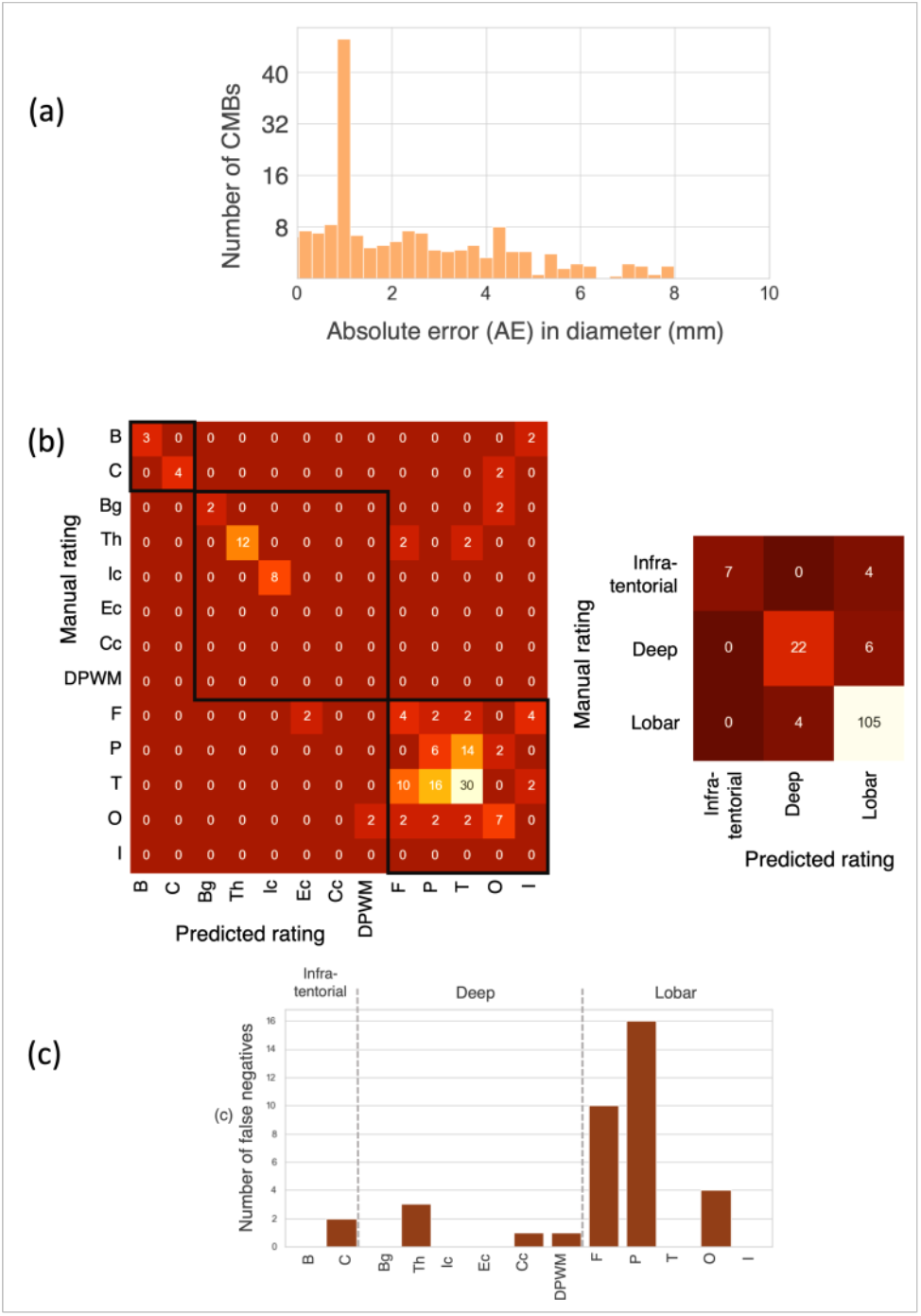
CMB size estimation and rating results on TICH2. (a) Histograms of absolute error (AE in mm) in the diameters between the manually segmented and the detected true positive (TP) CMBs. Confusion matrices for automated CMB rating for individual structures and regions using (b) detected TP CMBs, (c) bar plots showing the number of FNs for individual structures.

## 3 Results

Figure 2(a) shows the histogram of AE between diameters of manually segmented and predicted TP CMBs. Around 35% of detected TPs differed from the manually segmented CMBs by *<* 1.0 mm, and only 6% of CMBs (11 CMBs) had a difference *>* 6.0 mm. We obtained a mean absolute error (MAE) value of 2.0 mm between the manually segmented and predicted TP CMBs. Figure 2(b) shows the confusion matrices (both region- and structure-wise) between the manual and automated ratings obtained for predicted CMBs. Our automated MARS rating method achieved a region-wise sensitivity of 0.64, 0.79, 0.96, a region-wise precision of 1.00, 0.85, 0.91 and region-wise F1-measure of 0.78, 0.82, 0.93 for infratentorial, deep and lobar regions respectively, with an overall accuracy of 91%. Figure 2(c) shows the structure-wise distribution of FNs. From the figure, it is evident that most of FNs occurred in the parietal and frontal lobes. Table 1 reports the statistics of the subject-wise predicted CMB counts (including TPs and FPs) across all subjects.

## 4 Discussion

We extracted the size (diameter) of the CMBs and automatically calculated the MARS score. On comparing the CMB sizes, we observed that most of the cases with high AE (≈ 6mm) were due to the nearest small fragments of vessels that were segmented along with the CMBs. The MARS regions obtained using our method aligns well with those of clinically provided manual rating (indicated by a high performance of 91% overall accuracy). In our work, we have considered only true positive (TP) CMBs (since the confusion matrix requires manual ratings), however note that it is possible to obtain the counts of FPs also in the MARS regions, as indicated by total CMB counts (TPs and FPs) reported in table 1). We observed more FPs in the lobar regions (especially in the frontal, temporal and parietal lobes) indicating that the possible source of FPs could be the edge artefacts close to the skull and the troughs of the cortical sulci.

FNs were mostly the subtle and small CMBs, especially when they occur closer to the blood vessels. Overall, the count of CMBs in individual regions (reported in Table 1) can be used as location-based soft priors/weights for further reducing FPs and FNs and for improving the accuracy of automated CMB detection methods. The misclassifications occurred mainly at the structure-level, near their boundaries and could be due to registration errors, size of dilation kernels and outline errors occurring from the segmentation errors in predicted CMBs, especially in the region/structure boundaries (hence shifting the centroids to the adjacent regions). Regarding region-wise vs structure-wise accuracy, the performance measures obtained for the three main regions (infratentorial, deep, lobar) were highly robust, despite minor misclassifications across the individual structures (e.g., basal ganglia) (Figure 2). Clinically, clinical impacts of CMBs are studied often at the region-level - e.g., lobar CMBs are associated with CAA and Alzheimer’s disease, while deep CMBs are associated with cerebrovascular diseases [5, 20]. Given a high region-wise accuracy for CMB counting and accurate size determination, the proposed method could be promising in extracting imaging-based CMB biomarkers.

## 5 Conclusions

In this work, we proposed automated characterisation of CMBs based on their size and spatial distribution using MARS rating scale. Our method gave an MAE of 2.0 mm between diameters of manually segmented and predicted CMBs. For automated rating of CMB counts, we obtained an overall accuracy *>*90% for predicted CMBs, indicating the reliability of the atlas registration performed in our method. A future direction of this work will be to investigate the relationship between the region-wise distribution of CMBs and various clinical/demographic factors, to further study the impact of CMBs at population-level.

## Data Availability

The Python code for the implementation of the method and the atlas of MARS regions in MNI space are available at https://github.com/v-sundaresan/microbleed-size-counting.

## Abbreviations

AE: Absolute Error
BOMBS: Brain Observer Microbleed Scale
CAA: Cerebral Amyloid Angiopathy
CMB: Cerebral Microbleeds
DTI: Diffusion Tensor Imaging
FAST: FMRIB’s Automated Segmentation Tool
FLIRT: FMRIB’s Linear Image Registration Tool
FMRIB: Oxford Centre for Functional MRI of the Brain
FN: False Negatives
FSL: FMRIB Software Library
FP: False Positives
ICBM: International Consortium of Brain Mapping
JHU: Johns Hopkins University
MAE: Mean Absolute Error
MARS: International Consortium of Brain Mapping
MNI: Montreal Neurological Institute
MRI: Magnetic Resonance Imaging
SWI: Susceptibility-weighted Imaging
TICH2: Tranexamic acid for IntraCerebral Haemorrhage2 trial
TP: True Positives

## Declarations

### Ethics approval and consent to participate

The TICH-2 trial obtained ethical approval from East Midlands (Nottingham 2) NHS Research Ethics Committee (Reference: 12/EM/0369) and the amendment to allow the TICH2 MRI sub-study was approved in April 2015 (amendment number SA02/15). The patients/participants provided their written informed consent to participate in this study.

### Consent for publication

Not applicable

### Availability of Data and Material

The TICH-2 MRI sub-study data can be shared with bona fide researchers and research groups on written request to the sub-study PI RD. Proposals will be assessed by the PI (with advice from the TICH-2 trial Steering Committee if required) and a Data Transfer Agreement will be established before any data are shared.

### Competing interests

MJ and LG receive royalties from licensing of FSL to non-academic, commercial parties. The remaining authors declare that the research was conducted in the absence of any commercial or financial relationships that could be construed as a potential conflict of interest.

### Funding

This research was supported by the NIHR Oxford Health Biomedical Research Centre (NIHR203316). The views expressed are those of the author(s) and not necessarily those of the NIHR or the Department of Health and Social Care. The Wellcome Centre for Integrative Neuroimaging is supported by core funding from the Wellcome Trust (203139/Z/16/Z and 203139/A/16/Z). For the purpose of open access, the author has applied a CC-BY public copyright licence to any Author Accepted manuscript version arising from this submission. This work was also supported by the Engineering and Physical Sciences Research Council (EPSRC), Medical Research Council (MRC) (grant number EP/L016052/1) and NIHR Nottingham Biomedical Research Centre. The TICH-2 MRI sub-study was funded by a grant from British Heart Foundation (grant reference PG/14/96/31262) and the TICH-2 trial was funded by a grant from the NIHR Health Technology Assessment programme (project code 11 129 109). VS was supported by the Wellcome Centre for Integrative Neuroimaging (203139/Z/16/Z). VS is currently supported by DBT/Wellcome Trust India Alliance Fellowship [grant number IA/E/22/1/506763] and Pratiksha Trust, Bangalore, India [grant number FG/PTCH-23-1004]. GZ is supported by the Italian Ministry of Education (MIUR) and by a grant Dipartimenti di eccellenza 2018-2022, MIUR, Italy, to the Department of Biomedical, Metabolic and Neural Sciences, University of Modena and Reggio Emilia. PR is in receipt of a NIHR Senior Investigator award. MJ was supported by the NIHR Oxford Biomedical Research Centre (BRC), and this research was funded by the Wellcome Trust (215573/Z/19/Z). LG was supported by an Alzheimer’s Association Grant (AARF-21-846366) and by the NIHR Oxford Health Biomedical Research Centre (NIHR203316).

### Authors’ contributions

VS contributed to conceptualization, methodology, software, validation, formal analysis, investigation, visualization, and wrote the original draft. GZ, AM, RD, PR, DA, and NS contributed to resources. RD and SS contributed to data curation. DA contributed to data curation and investigation. LG contributed to conceptualization, methodology, data curation, and project administration. MJ contributed to conceptualization, methodology, funding acquisition, and project administration. GZ, MJ, and LG contributed to supervision. All authors contributed to manuscript revision, read, and approved the submitted version.

## Acknowledgements

The content of this manuscript is derived from the D.Phil. (Ph.D.) thesis of the first author (VS). The thesis titled “Detection of cerebral small vessel disease signs on brain MR images” (https://ora.ox.ac.uk/objects/uuid:10f3edb1-2566-4f0c-b4cb-0c581582c6c0) has been deposited in Oxford University Research Archive (ORA) as per the university guidelines. The authors confirm that the thesis content related to this manuscript has not publicized elsewhere in any form.

## Notes

### Competing Interest Statement

The authors have declared no competing interest.

### Author Declarations

The TICH-2 trial obtained ethical approval from East Midlands (Nottingham 2) NHS Research Ethics Committee (Reference: 12/EM/0369) and the amendment to allow the TICH2 MRI sub-study was approved in April 2015 (amendment number SA02/15). The patients/participants provided their written informed consent to participate in this study. The TICH-2 MRI sub-study data can be shared with bona fide researchers and research groups on written request to the sub-study PI RD. Proposals will be assessed by the PI (with advice from the TICH-2 trial Steering Committee if required) and a Data Transfer Agreement will be established before any data are shared. More details at https://www.thebottomline.org.uk/summaries/icm/tich-2/.

